# Estimation of SARS-CoV-2 infection fatality rate by real-time antibody screening of blood donors

**DOI:** 10.1101/2020.04.24.20075291

**Authors:** Christian Erikstrup, Christoffer Egeberg Hother, Ole Birger Vestager Pedersen, Kåre Mølbak, Robert Leo Skov, Dorte Kinggaard Holm, Susanne Sækmose, Anna Christine Nilsson, Patrick Terrence Brooks, Jens Kjærgaard Boldsen, Christina Mikkelsen, Mikkel Gybel-Brask, Erik Sørensen, Khoa Manh Dinh, Susan Mikkelsen, Bjarne Kuno Møller, Thure Haunstrup, Lene Harritshøj, Bitten Aagaard Jensen, Henrik Hjalgrim, Søren Thue Lillevang, Henrik Ullum

## Abstract

**Background:** The pandemic due to severe acute respiratory syndrome coronavirus 2 (SARS-CoV-2) has tremendous consequences for our societies. Knowledge of the seroprevalence of SARS-CoV-2 is needed to accurately monitor the spread of the epidemic and also to calculate the infection fatality rate (IFR). These measures may help the authorities to make informed decisions and adjust the current societal interventions. Blood donors comprise approximately 4.7% of the similarly aged population of Denmark and blood is donated in all areas of the country. The objective of this study was to perform real-time seroprevalence surveying among blood donors as a tool to estimate previous SARS-CoV-2 infections and the population based IFR.

**Methods:** All Danish blood donors aged 17-69 years giving blood April 6 to 17 were tested for SARS-CoV-2 immunoglobulin M and G antibodies using a commercial lateral flow test. Antibody status was compared between areas and an estimate of the IFR was calculated. The seroprevalence was adjusted for assay sensitivity and specificity taking the uncertainties of the test validation into account when reporting the 95% confidence intervals (CI).

**Results:** The first 9,496 blood donors were tested and a combined adjusted seroprevalence of 1.7% (CI: 0.9-2.3) was calculated. The seroprevalence differed across areas. Using available data on fatalities and population numbers a combined IFR in patients younger than 70 is estimated at 82 per 100,000 (CI: 59-154) infections.

**Conclusions:** The IFR was estimated to be slightly lower than previously reported from other countries not using seroprevalence data. The IFR, including only individuals with no comorbidity, is likely several fold lower than the current estimate. This may have implications for risk mitigation. We have initiated real-time nationwide anti-SARS-CoV-2 seroprevalence surveying of blood donations as a tool in monitoring the epidemic.

## Introduction

Humanity is suffering from a pandemic due to severe acute respiratory syndrome coronavirus 2 (SARS-CoV-2). The local severity of the epidemic and experiences from other countries are used by the health authorities to calibrate societal interventions. These interventions, e.g. the closing of schools, public institutions, prohibition of group gatherings, and even curfew, have tremendous consequences.

The authorities rely on accurate real-time data to make informed decisions. Thus, numbers of patients tested positive for SARS-CoV-2, admitted to hospital, needing respiratory assistance or deceased from coronavirus disease 2019 (COVID-19) are updated on a daily basis. In contrast, little information exists on the percentage of the population with previous mild or asymptomatic COVID-19. The proportion of the population who have overcome the infection can probably be approximated by testing for antibodies against SARS-CoV-2. Antibodies may confer immunity to repeat infection and a high proportion of immune individuals can attenuate the epidemic. Measures of anti-SARS-CoV-2 seroprevalence can also be used to estimate the clinical impact of COVID-19. Statistics on COVID-19 morbidity and mortality vary greatly due to varying testing strategies and e.g. the capacity of the health care system to treat infected patients^1^. Countries that diagnose mild infections will report lower morbidity and mortality compared to those with a less comprehensive testing strategy. An accurate measure of seroprevalence can be used to estimate the accumulated number of SARS-CoV-2 infections and thus the infection fatality rate (IFR) in the underlying population.

Blood donors comprise approximately 4.7% of the Danish population in the same age group^2^. Healthy volunteers donate blood in all areas of the country ensuring wide geographical coverage. We have initiated a prospective screening of all blood donations for SARS-CoV-2 antibodies to establish a real-time nationwide overview of antibody status. The objective of this study is to perform a seroprevalence survey among blood donors as a tool in the monitoring of the SARS-CoV-2 epidemic.

## Methods

In Denmark, approximately 270,000 blood donations are given annually. All Danish blood donation facilities participated in this survey. From April 6 to April 17, 2020 a total of 9,496 blood donations were given by 17–69-year-old donors. Blood donors are healthy and must comply with strict eligibility criteria^3^. Currently, donors must self-defer for two weeks if they develop fever with upper respiratory symptoms.

SARS-CoV-2 IgG and IgM antibodies were tested on EDTA plasma or whole blood by a lateral flow test according to the manufacturer’s recommendations (IgM/IgG Antibody to SARS-CoV-2 lateral flow test, Livzon Diagnostics Inc., Zhuhai, Guangdong, China). Samples were concluded as reactive if the IgM, the IgG, or both bands were visible. A validation of the lateral flow test was performed. A total of 651 plasma samples from blood donors giving blood before November 2019 were tested (3 reactive of 651 samples, 1 inconclusive). Specificity was estimated to be 99.54% (98.66-99.90). Samples from 155 patients with previous SARS-CoV-2 were tested; 128 were reactive. Sensitivity was thus estimated to be 82.58% (75.68-88.20). Inter-observer agreement was assessed by three independent raters of 299 samples. The observed Kappa (Fleiss) score was 92% for the combined rating of either IgM and/or IgG positivity. Validation and testing were performed by experienced staff in five regional blood establishments.

We retrieved data on population numbers as of January 1^st^ 2020^4^ and the number of infected and deceased due to COVID-19 using daily updated data^5^.

### Statistics

Statistical analysis was performed in RStudio 1.2 and R 3.6.0. Results were reported as percentages with 95% confidence intervals (CI). The EpiR package was used to adjust seroprevalence for sensitivity and specificity. We used the Rogan Gladen estimate to calculate the true prevalence. CI were derived by 10^8-sample percentile bootstrapping independently sampling sensitivity, specificity and apparent prevalence using binomial distributions.

### Ethics

SARS-CoV-2 antibody testing was performed as a routine screening of all blood donations. Only consenting donors were tested and informed about their result. Anonymized data was used in this study. The Regional Scientific Ethical Committees for the Zealand Region of Denmark approved the investigation as a register project (20-000013).

## Results

We included blood donors aged 17 to 69 years and a total of 9,496 blood donors were informed and all consented to testing; see Table 1 for characteristics.

**Table 1.** Age and sex stratified seroprevalence of anti-SARS-CoV-2.

| Age strata | Female |  |  | Male |  |  | Total |  |  |
| --- | --- | --- | --- | --- | --- | --- | --- | --- | --- |
|  | Non-reactive | Reactive |  | Non-reactive | Reactive |  | Non-reactive | Reactive |  |
|  | n | n | % | n | n | % | n | n | % |
| 17-29 | 1,477 | 33 | 2.2 | 901 | 28 | 3.0 | 2,378 | 61 | 2.5 |
| 30-39 | 838 | 5 | 0.6 | 914 | 11 | 1.2 | 1,752 | 16 | 0.9 |
| 40-49 | 1,067 | 25 | 2.3 | 1,062 | 21 | 1.9 | 2,129 | 46 | 2.1 |
| 50-59 | 953 | 8 | 0.8 | 1,082 | 18 | 1.6 | 2,035 | 26 | 1.3 |
| 60-69 | 478 | 11 | 2.2 | 551 | 13 | 2.3 | 1,029 | 24 | 2.3 |
| Total | 4,813 | 82 | 1.7 | 4,510 | 91 | 2.0 | 9,323 | 173 | 1.8 |

The distribution between seropositivity for IgM and IgG appears in Table 2.

The estimated number of infected individuals was calculated per area in the relevant age group (Table 3). The overall unadjusted seroprevalence was 1.8% (CI: 1.6-2.1). After adjusting for assay sensitivity and specificity including their CI, the overall seroprevalence was 1.7% (CI: 0.9-2.3).

**Table 2.** Samples stratified according to detectable SARS-CoV-2 IgM or IgG antibody isotype.

|  | Non-reactive |  |  |  |
| --- | --- | --- | --- | --- |
|  |  | IgM-only | IgG-only | IgM+IgG |
| n | 9,323 | 69 | 63 | 41 |
| % of tested | 98.2 | 0.73 | 0.66 | 0.43 |
| % of reactivities | - | 39.9 | 36.4 | 23.7 |

**Table 3.**
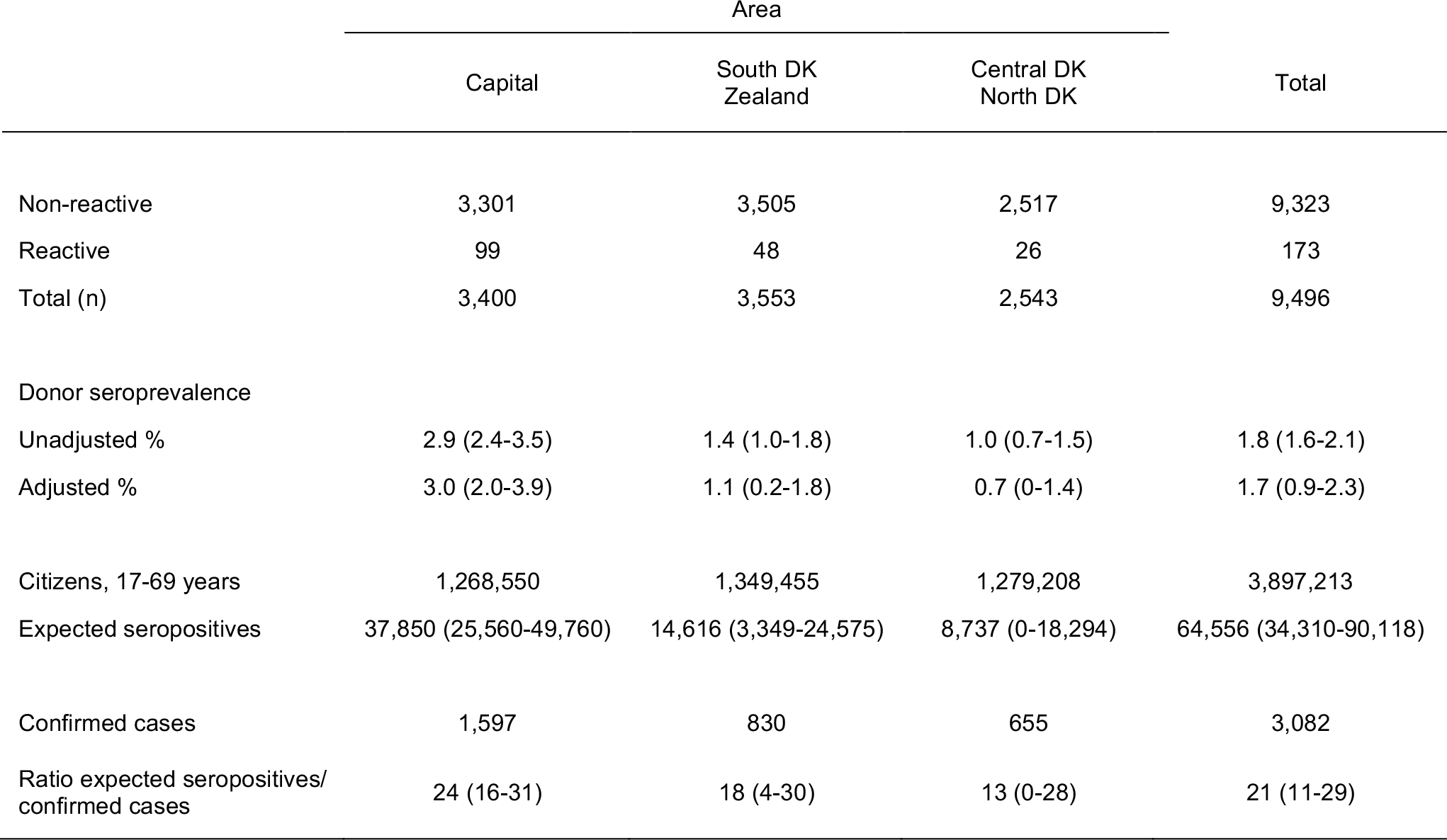
Distribution of seroprevalence according to area. A test sensitivity of 82.58% (75.68-88.20) and a specificity of 99.54% (98.66-99.90) was used in the adjustment of the seroprevalence percentage. Confirmed cases in each area were defined as confirmed viral RNA reactives as of April 1, 2020 to allow for an extra two-week lag time between case confirmation and antibody development.

The seroprevalence in the Capital Region was higher than in the other four regions combined (3.0% vs 0.9%, difference: 2.1 percentage points, CI: 1.3-2.9).

As of April 21, 2020, 370 individuals are reported to have died from SARS-CoV-2 in Denmark; 53 of these were younger than 70. Thus, the combined IFR in patients younger than 70 is estimated at 82 per 100,000 infections (CI: 59-154). The total ratio between estimated antibody-positive individuals and the number of confirmed cases was 21 (CI: 11-29).

## Discussion

In this survey of SARS-CoV-2 antibodies in Danish blood donors we found a seroprevalence of 1.7 (CI: 0.9-2.3) adjusted for the assay performance and a low IFR of 82/100,000 (CI: 59-154). This IFR of 0.082% is slightly lower than a recently published COVID-19 IFR estimate of 0.145% (CI: 0.088-0.317, individuals below 60 years) not including seroprevalence data^6^. The ratio between estimated antibody-positive individuals and confirmed COVID-19 cases is expected given the targeted early Danish SARS-CoV-2 testing strategy. The lack of large seroprevalence surveys prevents a comparison with other areas/countries. The low IFR is encouraging, but several caveats exist. Although blood donors represent a very broad population base, they are selected healthy and self-defer for two weeks after signs of COVID-19. Conversely, blood donor prevalence increases with income^7^ and we speculate that this leads to higher risk of exposure through travel and social activity. We may therefore either under or overestimate the true population immunity.

We validated the antibody assay primarily in individuals diagnosed with clinical COVID-19. If silent and mild infections lead to weaker antibody responses, we will underestimate the population immunity. Also, screening only for antibodies may underestimate the prevalence of infections, if cellular cytotoxicity is able to eradicate virally infected cells, as for SARS-CoV, before eliciting a humoral response^8^. Finally, this study only addresses the IFR in 17–69-year-old individuals. The IFR in other population strata, e.g. among individuals above 80 or with comorbidity is higher^6,9^. Currently, the governments in most countries are trying to balance the economic consequences of a societal lockdown against the risk of an uncontrolled epidemic. Our results underpin that social distancing in a healthy population predominately acts as a means to protect vulnerable individuals. It would be challenging to perform an unbiased seroprevalence survey in the background population. As blood donation facilities are located nationwide and operate continuously the screening is suited to monitor regional differences and temporal changes. With greater knowledge of the seroprevalence in other population strata the continued monitoring may also be used to effectively model the activity of the SARS-CoV-2 epidemic.

### Limitations

We undertook a validation and found a less than perfect sensitivity of 82.6% (75.7-88.2) when previous PCR-confirmed COVID-19 patients were tested. However, it is known that not all infected individuals produce antibodies. The specificity was acceptable at 99.5% (98.7-99.9) but leads to a low positive predictive value in low-prevalence areas.

We used a conservative method to estimate the confidence interval and thus took not only the sample variation but also the uncertainty in the sensitivity and specificity into account. This is necessary because we, unlike most diagnostic and screening tests, do not have a Gold Standard to confirm positive or negative results. The confidence interval for the regions with lowest antibody prevalence thus reached a lower limit seroprevalence of 0%.

The estimates for the IFR should allow for the lag time from infection to death. Based on current literature time from infection to death in non-survivors is 23-30 days^10,11^. Similarly, the lag time from infection to the detection of antibodies may be 16 days^10,12^. Donor self-deferral due to respiratory symptoms will add to the lag time for the detection of antibodies. We used the last available total of deceased citizens due to COVID-19 (April 21, 2020). Using earlier values would result in a lower IFR estimate while waiting for later death tolls would result in a higher IFR. The death toll among all citizens below 70 years was used even though only 16 of 53 deaths appeared among individuals with no comorbidity. This was chosen because the denominator included all citizens in the age strata, thus, also individuals with comorbidity. The IFR including only individuals with not comorbidity is thus likely several fold lower than the current estimate. Rapid tests are read by individuals and inter-observer variation often exist. Furthermore, there is uncertainty regarding cross reactivity of SARS-CoV-2 and other coronavirus antibodies. The results included in this article will be updated and freely accessible at http://www.bloddonor.dk/antisarscov2.

## Conclusion

Our results indicate that the IFR among individuals aged 17 to 69 years is 82/100,000 (CI: 59-154). This may have implications for risk mitigation. The IFR in older population strata may be considerably higher. Nationwide continuous seroprevalence surveying of blood donations may be a tool in monitoring the SARS-CoV-2 epidemic.

## Data Availability

Aggregated data are available. Please see provided URL.

## Acknowledgements

We thank laboratory technicians from the Departments of Clinical Immunology at Aarhus University Hospital, Copenhagen University Hospital, Odense University Hospital, Zealand University Hospital, and Aalborg University Hospital for their excellent work during the validation of the assay and testing of the samples.

